# Fixed-Dose Ivermectin for Mass Drug Administration: Is it time to leave the dose pole behind? Insights from an Individual Participant Data Meta-Analysis

**DOI:** 10.1101/2025.04.16.25325920

**Authors:** Adriana Echazu, Daniela Bonanno, Pedro Emanuel Fleitas, Julie Jacobson, Gabriela Matamoros, Charles Mwandawiro, Wendemagegn Enbiale, Áuria de Jesus, Alan Brooks, Alejandro Javier Krolewiecki

**Affiliations:** Fundación Mundo Sano, Madrid, Spain; Instituto de Investigaciones de Enfermedades Tropicales (IIET), Universidad Nacional de Salta, Oran, Salta, Argentina; Instituto de Salud Global de Barcelona (ISGlobal), Barcelona, Spain; Bridges to Development, Switzerland & Unites States; Instituto de Investigaciones en Microbiología (IIM), Facultad de Ciencias, Universidad Nacional Autonoma de Honduras, Tegucigalpa, Honduras; Kenya Medical Research Institute (KEMRI), Nairobi, Kenya; Dermatovenerology Department, College of Medicine and Health Science, Bahir Dar University, Bahir Dar, Ethiopia; Centro de Investigação em Saúde de Manhiça (CISM), Manhiça, Maputo, Mozambique

## Abstract

**Background:** Ivermectin (IVM) is widely used in mass drug administration (MDA) programs for the control of neglected tropical diseases (NTDs). Current regimens rely on weight- or height-based dosing, which lead to operative challenges. This study evaluates an age-based fixed-dose regimen for IVM.

**Methodology:** This is an individual participant data (IPD) meta-analysis including anthropometric data from over 700,000 individuals, across 53 NTD-endemic countries. Fixed-dose regimens were developed based on weight distribution by age. The proportion of individuals achieving the target range dose (200–400 µg/kg) was assessed and compared to traditional dosing regimens.

**Principal Findings:** Fixed-doses of 3 mg for pre-school children (PSAC), 9 mg for school-aged children (SAC), and 18 mg for women of reproductive age (WRA) resulted in a higher proportion of participants receiving the target dose compared to weight- and height-based regimens (79.9% vs. 32.7% and 37.3%, respectively, p < 0.001). Underdosed individuals were fewer with fixed-dose (8.7%) compared to weight-based (32.6%) and height-based (46.3%) regimens. Although doses above the target range increased slightly, most remained within 600 µg/kg.

**Conclusions:** An age-based fixed-dose regimen for IVM could improve treatment coverage and simplify MDA activities. Simplified logistics could lead to cost savings in drug distribution and administration, improving the overall efficiency of MDA programs. These findings support the inclusion of currently excluded PSAC in IVM-based MDA interventions. More broadly, this paper provides evidence for considering the potential policy and programmatic implications of fixed-dose IVM. This Individual Participant Data Meta-analysis (IPD-MA) is registered in PROSPERO (CRD42024521610).

**AUTHOR SUMMARY:** Ivermectin is an essential drug with proven safety and effectiveness against several parasitic infections. It plays a key role in Mass Drug Administration (MDA) programs targeting prevalent Neglected Tropical Diseases. Currently, ivermectin dosing is based on weight or height, which can be difficult to measure in the field during MDA campaigns and adds complexity and workload for health workers. These methods also carry a risk of underdosing.

In this study, we analyzed data from more than 700,000 participants across 53 countries to explore whether a fixed-dose approach could simplify MDA implementation while maintaining doses within the therapeutic range. We found that fixed-dose regimens provide more accurate treatment for a larger proportion of individuals, reduce the likelihood of underdosing, and only occasionally result in doses above the recommended levels, typically by small margins and in a limited proportion of participants. This simplified approach could ease treatment delivery in community settings and improve coverage and operational efficiency. Our findings provide practical evidence to inform policy discussions on how to streamline and strengthen ivermectin - based MDA programs.

## INTRODUCTION

The control of several neglected tropical diseases (NTD) is based on the provision of single-dose anthelmintic drugs through mass drug administration (MDA) campaigns [1,2]. Ivermectin (IVM) is an essential medicine widely used in MDA activities [3]. It is the drug of choice against onchocerciasis, scabies and strongyloidiasis, and in combination with other anthelmintics, against lymphatic filariasis and trichuriasis [4–7].

IVM is typically administered using a weight-based dosing regimen, with the currently recommended dose being 200 µgr/kg of body weight. In MDA programs, the World Health Organization (WHO) recommends using height as a proxy for weight, implemented through a dose pole to facilitate large-scale delivery [1,8]. Under this approach, children shorter than 90 cm or weighing less than 15 kg are excluded from treatment. Early clinical trials evaluating the safety and efficacy of ivermectin for onchocerciasis demonstrated that single doses as high as 800 µgr/kg were as safe as lower doses [9]. Notably, while these studies support the safety of higher doses, a clear upper safety limit for ivermectin in humans has not yet been formally established.

Current IVM dosing strategies pose operational challenges in large-scale MDA programs. Several studies have shown that height or weight-based dosing often leads to sub-therapeutic treatment in certain populations [10,11]. Additionally, children appear to have an accelerated clearance of IVM, potentially requiring higher doses [12]. Despite its favourable safety profile, data on IVM use in children under 5 years of age remain limited. Nevertheless, a systematic review including 1088 children under 15kg of body-weight that received IVM for a variety of indications suggests a safety profile comparable to that observed in older children [13]. This evidence coupled with accumulating information on the safety of IVM at doses several times higher than currently recommended, supports the exploration of alternative fixed-dose regimens [14–16]. The European Medicines Agency’s Positive Opinion supporting age-based fixed-dosing of 9 and 18 mg of IVM in combination with albendazole aligns with this broader safety margin for IVM [17].

With over 500 million people receiving IVM annually through MDAs alone, the potential public health benefits of a fixed-dose approach are substantial [18,19]. This analysis aims to identify an alternative age-based dosing regimen using anthropometric data to evaluate fixed-dose regimens and compare drug exposure across current and exploratory strategies.

## METHODS

### Study design and registration

This is an Individual Participant Data Meta-analysis (IPD MA). The protocol has been registered at the International prospective register of systematic reviews PROPSERO: (PROSPERO 2024 CRD42024521610): https://www.crd.york.ac.uk/prospero/display_record.php?ID=CRD42024521610.

### Eligibility criteria

Studies were eligible if they included individual-level data on participants’ anthropometric measurements and the location.

Study-Level Inclusion Criteria:

1. Study design: Observational studies, public health surveys, and clinical trials.
2. Geographic scope: Countries or sub-national districts where preventive chemotherapy for STH is recommended according to WHO guidelines [1,20].
3. Time frame: Studies conducted between January 1, 2010, and December 31, 2024.
4. Data availability: Studies including anthropometric data. Participants with missing or incomplete IPD for any mandatory variable (country, age, sex, and weight) were excluded. Individuals with missing height data were retained, as height was considered non-essential.
5. Timing of data collection: Only baseline data was included.

Study-Level Exclusion Criteria:

1. Studies providing only aggregated data.
2. Studies enrolling severely ill subjects, such as those focused on tuberculosis or severe malaria.

Individual-Level Inclusion Criteria:

1. Age Groups: pre-school age children (PSAC): 2 to 4 years (24–59 months); school age children (SAC): 5 to 15 years; woman of reproductive age (WRA): 15 to 49 years.
2. Sex: Male and female (PSAC and SAC); only female adults (WRA) [1].

### IPD collection process and data integrity

IPD were obtained from two main sources: (1) studies on STH interventions conducted in endemic regions and (2) datasets from data repositories, accessed in compliance with their specific protocols.

The datasets underwent a five-step process: selection, standardization, compilation, cleaning, and consistency assessment. First, only variables of interest were retained, excluding others from the original studies. Next, standardization ensured consistency across datasets: age was recorded in months (children) or years (adults), weight in kilograms (one decimal), and height in centimeters (one decimal). Each site was coded by country.

This study assumed population homogeneity, justifying a one-stage meta-analysis, and during the compilation step, IPD from different studies were merged into three datasets: PSAC, SAC, and WRA. Cleaning followed, removing subjects with missing data. Finally, a consistency assessment was conducted using WHO Anthro (version 3.2.2) and WHO Anthro Plus (version 1.0.4) to detect potential measurement errors. Data were flagged as inconsistent if z-scores exceeded predefined thresholds:

- PSAC: WHZ < -5 or > 5; WAZ < -6 or > 5; HAZ < -6 or > 6; BAZ < -5 or > 5.
- SAC & WRA: WAZ < -6 or > 5; HAZ < -6 or > 6; BAZ < -5 or > 5.

These cut-offs, set by the software, were applied to identify extreme values likely resulting from measurement errors while accounting for diverse nutritional scenarios [21].

### Risk of bias assessment

The IPD compiled for this study were collected regardless of the design or outcomes of the original studies. Only baseline raw anthropometric IPD were included, rendering risks of bias related to outcome reporting or randomization processes irrelevant. Data completeness and consistency across studies were assessed, and participants with missing or inconsistent IPD were excluded according to pre-defined criteria applied systematically across all datasets. Since weight and height measurements were not conducted by our team, potential measurement errors, inaccuracies, or data entry mistakes could not be prevented. To minimize measurement bias, data flagged as inconsistent by the software were excluded. However, this approach may introduce selection bias if exclusions were not proportionally distributed across sites or other characteristics. The potential impact of these exclusions on the representativeness of the sample was considered in the analysis.

### Statistical analysis

The IPD were compiled into a database using Microsoft Excel (Microsoft, Redmond, WA). Data analysis was conducted with R version 3.1.1 (The R Foundation for Statistical Computing, GNU General Public License). Heterogeneity analysis was performed with the ‘metafor’ package [22]. Graphics were generated using the ‘ggprism’ extension of the ‘ggplot2’ package [23]. Map was created using QGIS version 3.43.10 (QGIS Development Team, released under the GNU General Public License, Version 2).

We applied a random effects model to address potential residual heterogeneity across the datasets included in the analysis and to examine the variation in effects across different subgroups. We assessed statistical heterogeneity using the I² statistic and τ², which allowed us to quantify the degree of variability.

Nutritional status of children was assessed using stunting proportions with 95% confidence intervals (CI) by country. Stunting was defined as a Height-for-Age Z-score (HAZ) below -2 SD from the international reference median. HAZ was selected as it applies to children from birth to 19 years. HAZ is also the best indicator of chronic malnutrition, often linked to STH infections. Malnutrition severity was classified by stunting prevalence using WHO thresholds: <20% (mild), 20–29% (moderate), 30–39% (high), and >40% (very high) [24].

The IPD were analyzed through a sequential process for the exploration of IVM dosing regimens as follows:

1. Selection of age-based fixed-dose The distribution of weight was analyzed to assess its dispersion. Each participant’s weight was multiplied by 200 to determine the exact IVM dose required to achieve the recommended 200 µg/kg. The median IVM dose was calculated for each year of age. Based on this median, the fixed-dose for each target group was determined by selecting the number of standard 3 mg tablets closest to the median dose required for the upper age limit of that group.
2. Dose Calculation The dose of IVM (in µg/kg) for each participant was calculated using three methods: i) Weight-based; ii) Height-based (dose-pole); iii) Fixed-dose as determined in Step 1. The weight and height- based doses used were those recommended by WHO [1,12].

3. Dose Categorization Participants were categorized into five groups based on the dose received under each regimen:

| IVM dosing regimen | Not currently recommended | 3 mg | 6 mg | 9 mg | 12 mg | 15 mg |
| --- | --- | --- | --- | --- | --- | --- |
| Weight based | < 15 Kg | 15 – 24 kg | 25 – 35 kg | 36 – 50 kg | 51 – 65 kg | 66 – 79 kg |
| Height based | < 90 cm | 90 – 119 cm | 120 – 139 cm | 140 – 159 cm | >159 cm |  |

- Not currently recommended: For weight-based regimen, body weight <15 kg; for height-based regimen, height <90 cm.
- Underdosed: <200 µg/kg.
- Target Range: 200–400 µg/kg.
- Above the Target Range (Level 1): 401–600 µg/kg.
- Above the Target Range (Level 2): 601–800 µg/kg.
- Above the Target Range (Level 3): > 800 µg/kg.
4. Comparison Between Regimens The proportion of participants in each dosing category (not currently recommended, undertreated, correctly treated, or above the target range) was calculated for each treatment regimen. To assess differences in proportions across regimens, pairwise comparisons with Chi-square test (α = 0.05) was performed. Additionally, median doses across the three regimens were compared using the Related Samples Friedman Test, stratified by group (PSAC, SAC, WRA). If significant differences were found, Wilcoxon signed-rank tests with Bonferroni correction were applied for post-hoc pairwise comparisons.

### Outcomes

The primary outcome of this study was: proportion of participants receiving target range dose of IVM (200 to 400 µg/kg) with a fixed-dose regimen based on age. A secondary outcome was to compare the performance of the fixed-dose regimen with height or weight-based dosing regimens. The measure of effect for this outcome were proportion and median doses differences between dosing regimens.

Prevalence of undernutrition in children globally and by country in the study population was another secondary outcome.

### Ethics Statement

This retrospective observational study did not require ethical approval, as it involved the analysis of de-identified data from previously conducted studies. All data were fully anonymized, ensuring that individual participants could not be identified and that confidentiality was maintained. The study relied on simulated ivermectin doses applied to hypothetical scenarios; no actual medication was administered, eliminating risks to any participants. Additionally, the study was conducted in accordance with established ethical standards, including the guidelines of the Council for International Organizations of Medical Sciences (CIOMS) in collaboration with the WHO, and the principles outlined in the Declaration of Helsinki [25].

## RESULTS

### IPD database sources

IPD were gathered from various original study datasets through the following sources: 1) Three studies focused on soil-transmitted helminths (STH) interventions conducted in Africa and Latin America [15,16,26]; 2) One study accessed via the Harvard Dataverse data repository [27]; 3) One dataset downloaded from the Digital Commons@Becker data repository [10]; 4) Datasets from 48 countries were requested to the Demographic and Health Surveys (DHS) Program [28]; and 5) Datasets from 62 studies were requested from the Infectious Diseases Data Observatory (IDDO)[29]. Participant flow diagram is described in Figure 1. During the data checking process, we identified some issues with the IPD: missing values for age or weight, inconsistent weight or height measurements, and formatting errors in age registration. These issues were addressed by excluding participants with any of these problems from the dataset. Exclusions were performed randomly based on predefined criteria, and no significant differences were observed in the distribution of excluded participants across countries. After the selection process, a total of 741,700 participants from 53 countries were included. The countries where preventive chemotherapy is recommended by WHO and those with available data to be represented in the study are highlighted in Figure 2.

**Figure 1:**
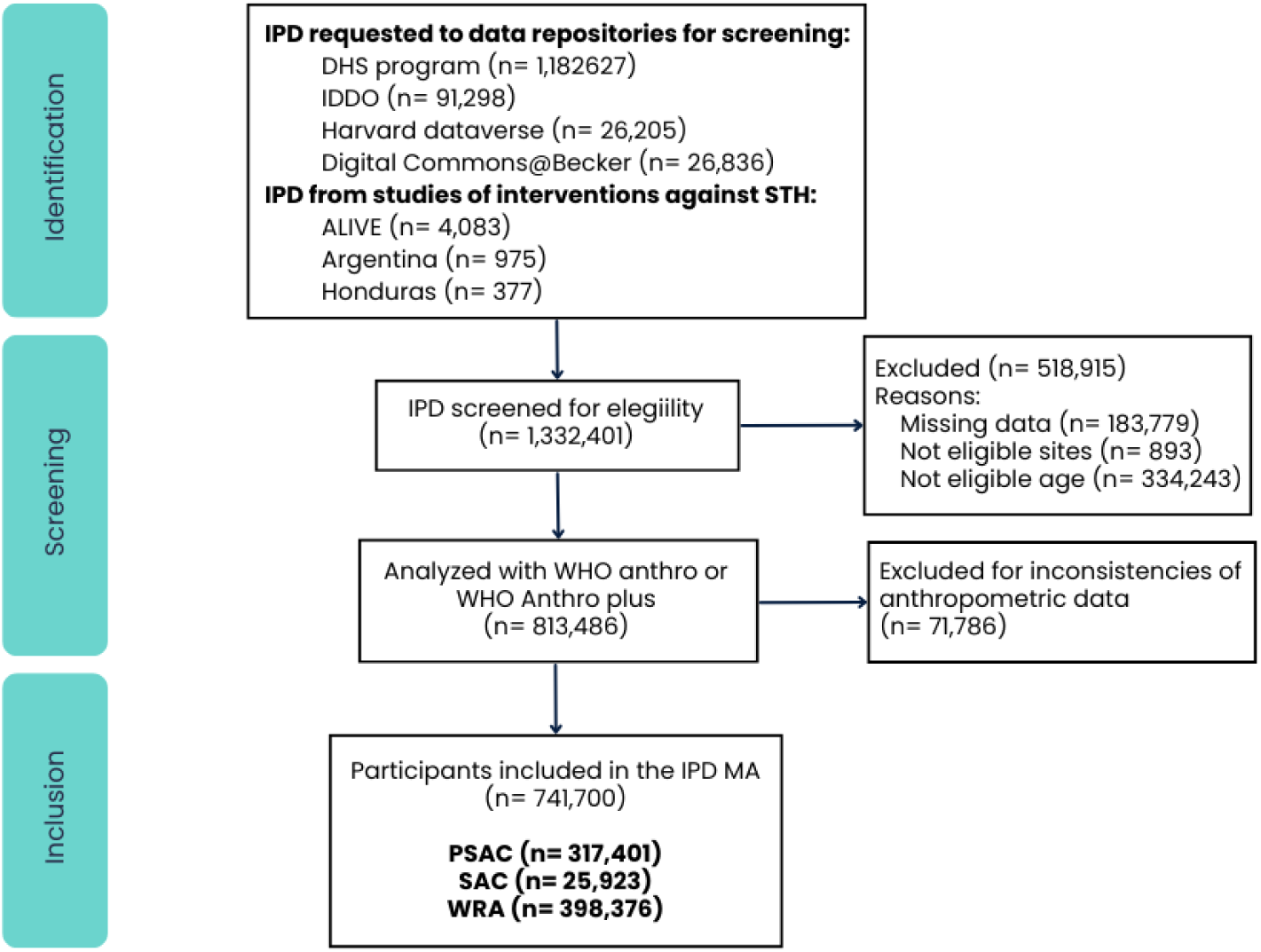
Flow diagram of participants of the Individual participant data meta-analysis of IVM fixed-dose feasibility.

**Figure 2:**
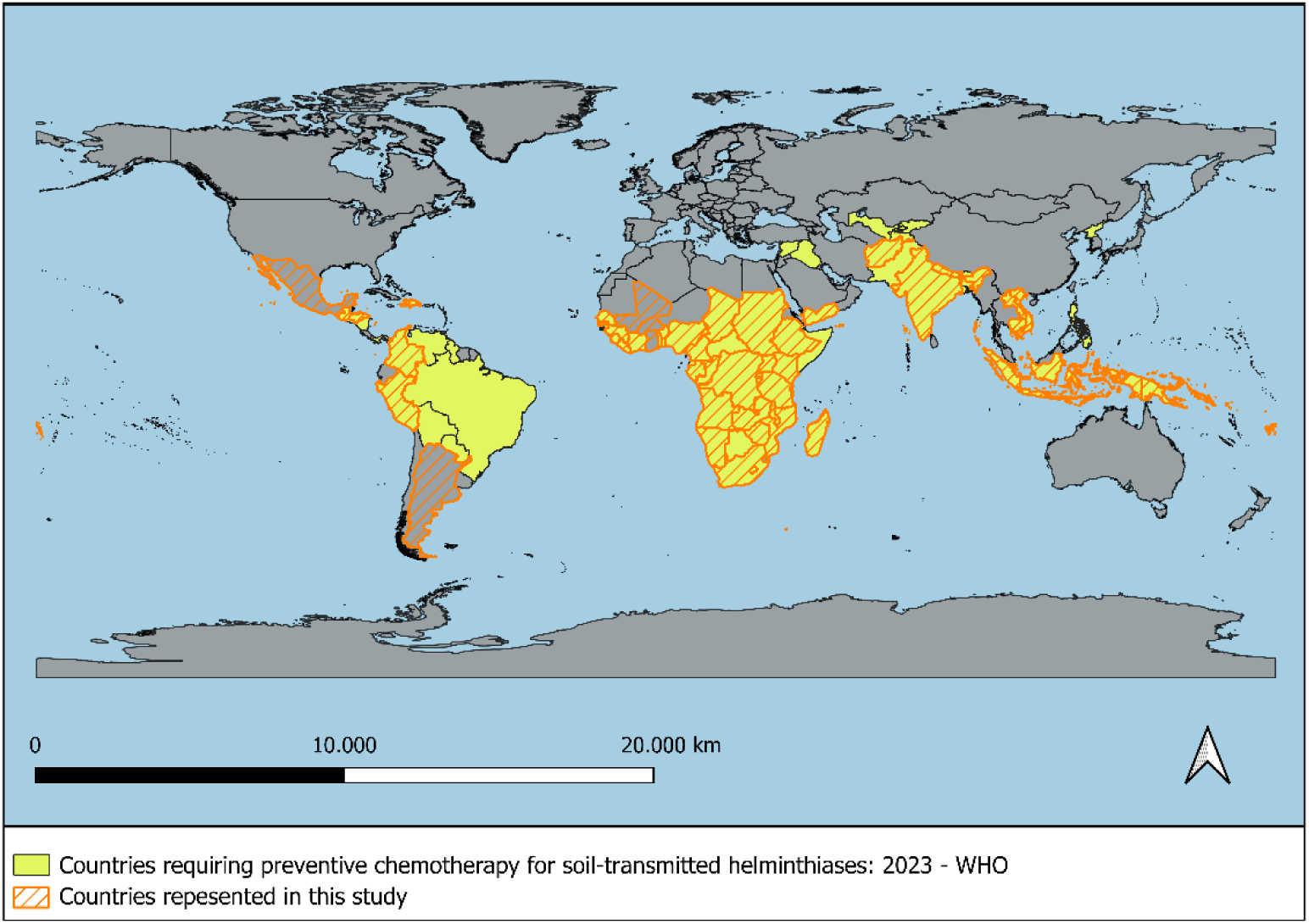
Map showing the countries included in the Individual Participant Data Meta-analysis.

### Characteristics of the study population

The study population included 398,376 WRA and 343,324 children, of whom 173,205 (50.4%; 95% CI: 50.3–50.6) were female. Among children, 317,401 (92.4%; 95% CI: 92.3–92.5) were PSAC, while 25,923 (7.55%; 95% CI: 7.4–7.6) were SAC. Heterogeneity analysis showed I² values of 17%, 18%, and 15% for PSAC, SAC, and WRA, respectively, indicating minimal heterogeneity. τ² values (0.005, 0.01, and 0.01) further confirmed low variability, suggesting that despite structural differences, heterogeneity across datasets was minimal.

Nutritional status assessment revealed that 120,718 children (35.2%; 95% CI: 35–35.3) were stunted (HAZ <-2). Consequently, the severity of malnutrition, as indicated by stunting prevalence, was classified as high across all countries (S1 table, Supporting information) [24].

### Ivermectin dose exploration

#### Fixed-dose Selection

The distribution of weight by age was analyzed, revealing a homogeneous sample. Among PSAC, the mean weight was 12.93 (SD: 2.45) kg, with a median of 12.70 kg (IQR: 11.2–14.4). In SAC, the mean was 27.25 (SD: 11.08) kg, and the median 24.80 kg (IQR: 19–33). For WRA, the mean was 54.99 (SD: 11.96) kg, and the median 52.90 kg (IQR: 46.8–60.9).

The weight-based calculation identified a median IVM dose of 2.54 mg (IQR: 2.24–2.88) for PSAC; 4.96 mg (IQR: 3.8–6.6) for SAC; and 10.58 mg (IQR: 9.36–12.18) for WRA, required to achieve the dose of 200 µg/kg. Percentiles of the calculated IVM dose by year of age for children and by group for WRA are presented in Table 1 (also figure S2 in the Supporting information).

**Table 1:** Percentiles of calculated amount of IVM required for reaching the recommended dose (200 µg/kg) by age.

| Target Group | AGE (years) | 5th Percentile (mg) | 10th Percentile (mg) | 25th Percentile (mg) | 50th Percentile (mg) | 75th Percentile (mg) | 90th Percentile (mg) | 95th Percentile (mg) |
| --- | --- | --- | --- | --- | --- | --- | --- | --- |
| PSAC | 2 | 1.72 | 1.82 | 2.02 | 2.22 | 2.46 | 2.70 | 2.88 |
|  | 3 | 2.00 | 2.10 | 2.32 | 2.56 | 2.84 | 3.10 | 3.30 |
|  | 4 | 2.24 | 2.40 | 2.60 | 2.88 | 3.18 | 3.50 | 3.72 |
| SAC | 5 | 2.40 | 2.80 | 3.00 | 3.20 | 3.60 | 4.00 | 4.40 |
|  | 6 | 2.80 | 2.80 | 3.20 | 3.60 | 4.00 | 4.63 | 5.00 |
|  | 7 | 3.00 | 3.06 | 3.43 | 4.00 | 4.52 | 5.05 | 5.60 |
|  | 8 | 3.20 | 3.40 | 3.90 | 4.40 | 5.00 | 5.61 | 6.20 |
|  | 9 | 3.42 | 3.80 | 4.20 | 4.80 | 5.60 | 6.26 | 7.00 |
|  | 10 | 3.80 | 4.00 | 4.60 | 5.20 | 6.00 | 7.00 | 7.80 |
|  | 11 | 4.20 | 4.60 | 5.03 | 6.00 | 6.87 | 8.00 | 9.00 |
|  | 12 | 4.40 | 4.80 | 5.20 | 6.12 | 7.40 | 8.80 | 10.00 |
|  | 13 | 5.00 | 5.41 | 6.20 | 7.28 | 8.60 | 10.20 | 11.39 |
|  | 14 | 5.53 | 6.00 | 7.00 | 8.00 | 9.40 | 10.94 | 12.30 |
|  | 15 | 6.20 | 6.80 | 7.60 | 8.80 | 10.00 | 11.60 | 12.94 |
| WRA | 15–45 | 7.94 | 8.42 | 9.36 | 10.58 | 12.18 | 14.06 | 15.46 |

Note: Dose calculated for each participant with the formula: weight (kg) \* 0.2 mg.

Based on these findings, fixed-dose regimens of 3 mg for PSAC and 9 mg for SAC were chosen to approximate the median required dose for the upper age limit of each group of children (median value for 4 yo for PSAC, median value for 15 yo for SAC). For WRA, an 18 mg fixed -dose was selected to approximate the 95th percentile for this group, minimizing the risk of underdosing. The median dose of IVM with the selected fixed-dose regimen by age group was 236 µg/kg (IQR: 208–267) for PSAC; 363 µg/kg (IQR: 272–473) for SAC and; 340 µg/kg (IQR: 295–384) for WRA. The highest variability observed was in the SAC group. Figure 3 Panel A displays the distribution of IVM dose by age group using the fixed-dose regimen with median and IQR.

**Figure 3:**
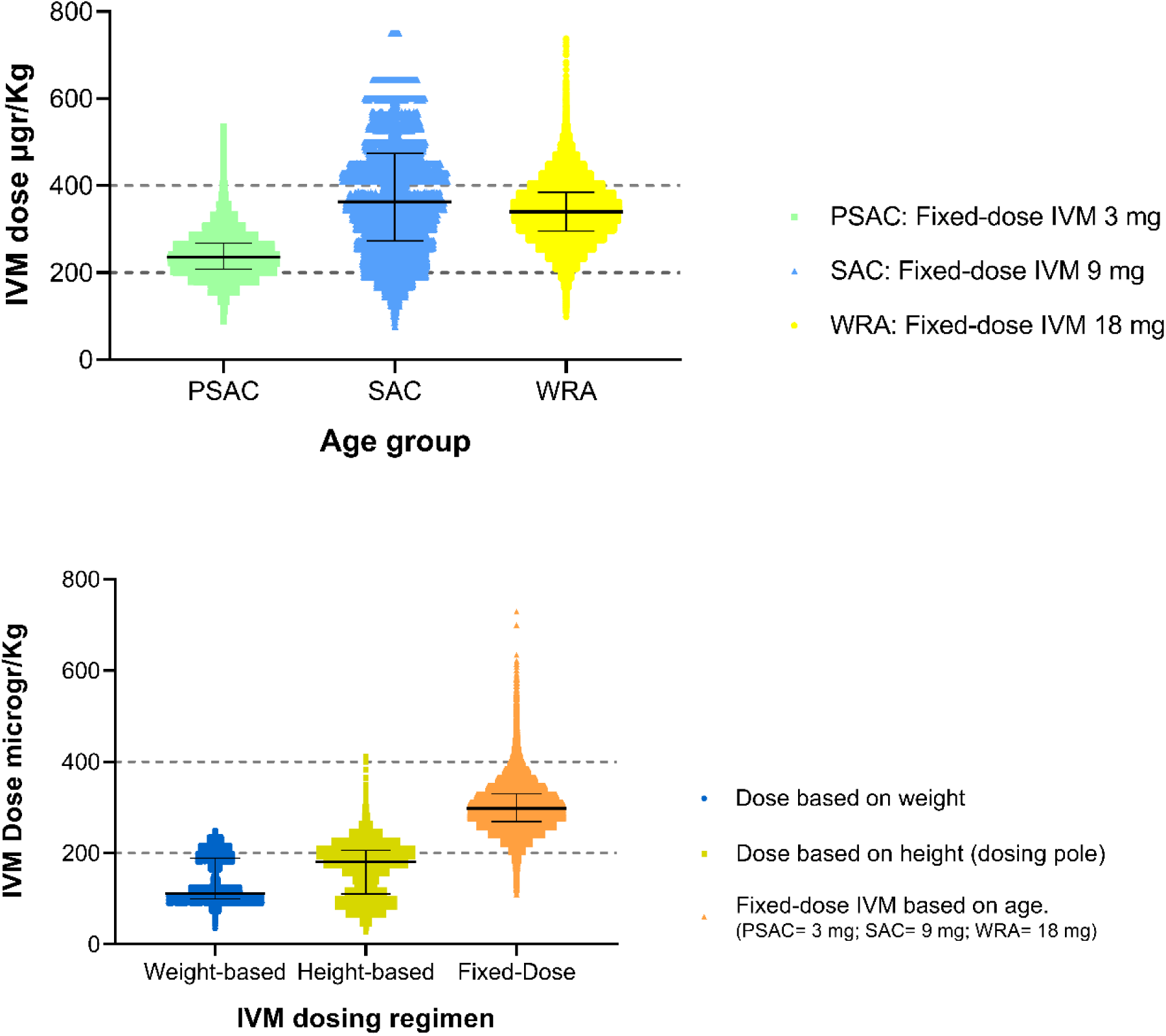
Ivermectin dose (µg/kg) distribution across dosing regimens. Panel A: Ivermectin dose using the fixed-dose regimen by age group (PSAC, SAC, and WRA); Panel B: Distribution of ivermectin doses across the study population, comparing weight-based, height-based, and fixed-dose regimens.

### Comparison Between Dosing Regimens

Considering the entire study population (PSAC, SAC, and WRA), the fixed-dose regimen resulted in the highest proportion of participants receiving the target range dose (79.9%), compared to the weight-based (32.7%) and height-based regimens (37.3%), (p<0.001). Conversely, the fixed-dose regimen had the lowest proportion of participants classified as underdosed (8.7%), with this difference also being statistically significant compared to the other regimens (p<0.001) (Table 2). The proportion of participants classified as receiving doses above the recommended range was higher with the fixed-dose regimen (11.27%), a difference that was statistically significant compared to the other regimens. However, only 0.02% of participants in this group exceeded the highest dose threshold (>800 µg/kg, level 3), reaching a maximum dose of 1125 µg/kg. The large majority of those receiving doses above the target range remained within level 1 (400–600 µg/kg). Figure 3 Panel B illustrates the distribution of IVM doses across the study population, presenting the median and interquartile ranges for each dosing approach, with most participants receiving doses below the target range of 200–400 µg/kg with both weight-based and height-based regimens. Figure 4 displays the proportion of participants by dose category and by dosing regimen for each age group.

**Figure 4:**
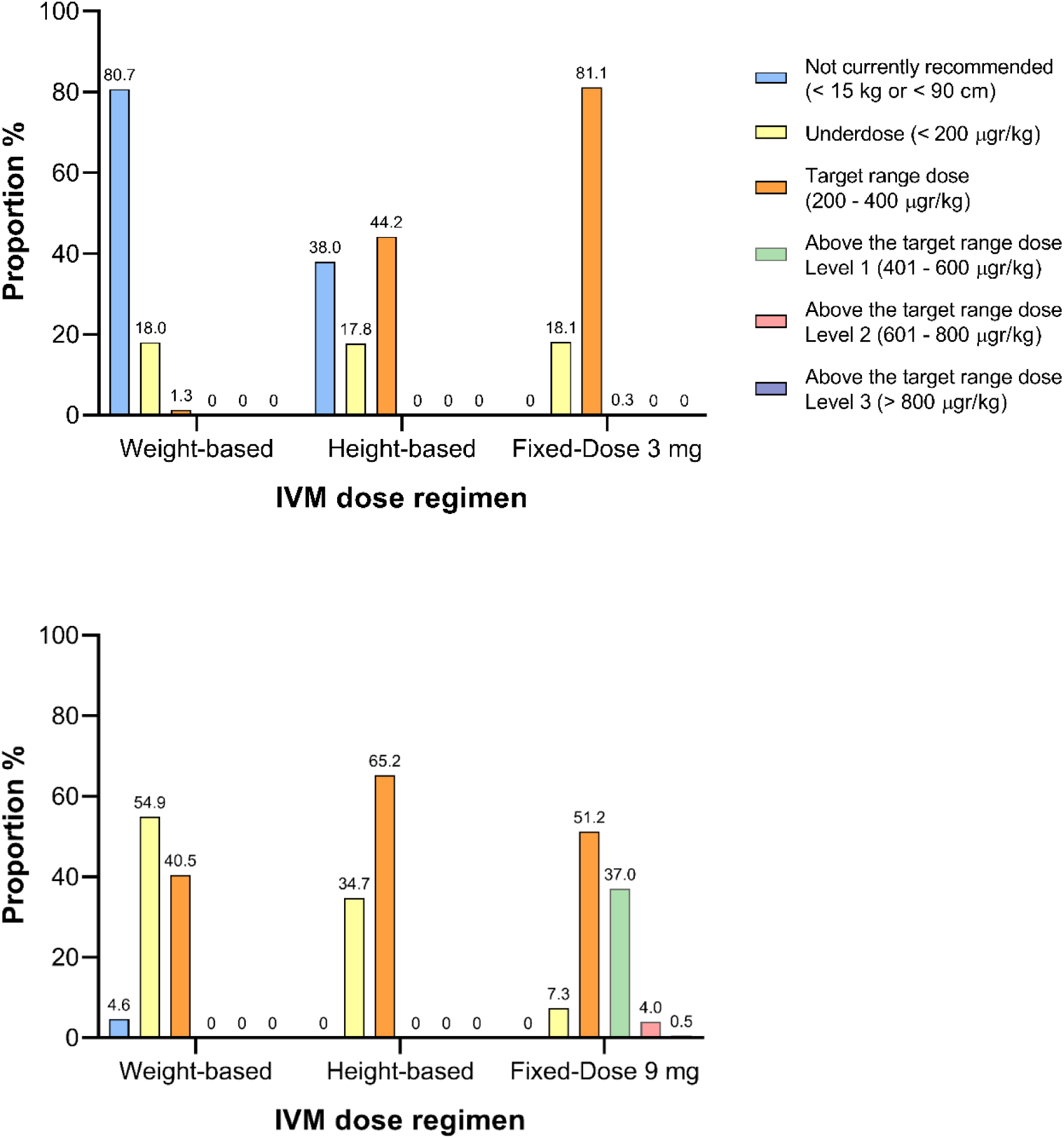
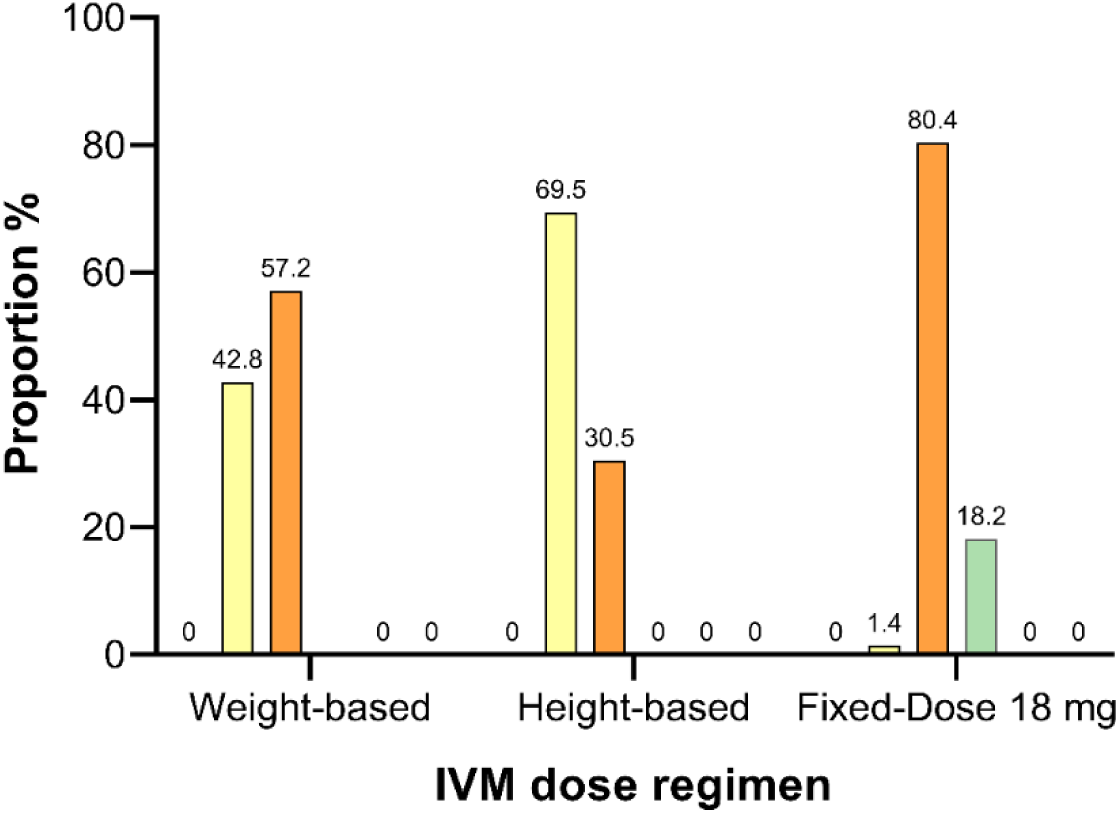
Proportion of participants by dose category and by dosing regimen for each age group. Panel A: Pre-school age children (PSAC); Panel B: School age children (SAC); Panel C: Woman of reproductive age (WRA).

**Table 2:** Proportion of participants within each dosing category by dosing regimen (N= 741.700)

| Dose Classification | Weight-Based (WB) % (95% CI) | Height-Based (HB) % (95% CI) | Fixed-dose (FD) % (95% CI) | p-value<br>p1: WB vs HB<br>p2: WB vs FD<br>p3: HB vs FD |
| --- | --- | --- | --- | --- |
| Not currently recommended * | 34.7 (34.6 – 34.8) | 16.4 (16.3 – 16.5) | NA | p1: <0.001 |
| Underdosed (< 200 µgr/kg) | 32.6 (32.5 – 32.7) | 46.3 (46.2 – 46.5) | 8.7 (8.6 – 8.8) | p1; p2; p3: <0.001 |
| Target Range Dose (200 – 400 µgr/kg) | 32.7 (32.5 – 32.8) | 37.3 (37.2 – 37.4) | 79.9 (79.8 – 80) | p1; p2; p3: < .001 |
| Above the Target Range Dose Level 1 (401 – 600 µgr/kg) | 0 | 0.003 (0.002 – 0.004) | 11.1 (11 – 11.2) | p1; p2; p3: <0.001 |
| Above the Target Range Dose Level 2 (601 – 800 µgr/kg) | 0 | 0 | 0.15 (0.14 – 0.16) | p1: 1 - p2; p3: <0.001 |
| Above the Target Range Dose Level 3 (< 800 µgr/kg) | 0 | 0 | 0.02 (0.016 – 0.022) | p1: 1 - p2; p3: <0.001 |

Notes: \* Not currently recommended: For weight-based regimen, body weight <15 kg; for height-based regimen, height <90 cm. NA: No exclusion criteria were applied for the age-based fixed-dose regimen.

Differences in the proportion of participants receiving the target IVM dose with the fixed-dose regimen were analyzed across countries. No significant variation was found between countries with a high prevalence of malnutrition and those without. Moreover, countries with severe stunting had fewer underdosed participants with fixed-dose compared to other countries.

In the comparison of median IVM doses, the fixed-dose regimen (median dose: 298; IQR: 269–330 µgr/kg) consistently provided a higher dose compared to the weight-based (median dose: 112; IQR: 99.5–189) µgr/kg) and height-based (median dose: 181; IQR: 110–206 µgr/kg) regimens. Additionally, current regimens failed to achieve the intended 200 µg/kg dose. In contrast, the fixed-dose regimen consistently maintained doses within the target range. In PSAC, the fixed-dose regimen provided a higher dose than both the height-based (Z = -321.021, p<0.001) and weight-based regimens (Z = -438.539, p < 0.001). In SAC, the fixed-dose regimen yielded a higher dose than the height-based (Z = -63.915, p < 0.001) and weight-based regimens (Z = -126.721, p <0.001). In WRA, the fixed-dose regimen again provided a higher dose compared to both the height-based (Z = -245.987, p < 0.001) and weight-based regimens (Z = -546.611, p<0.001). Since all comparisons yielded consistently low p-values (p<0.001), Bonferroni correction was not required.

### PSAC subgroup analysis

Among PSAC, 80.7% (95% CI: 80.6–80.9) of children were excluded from IVM use based on the weight-based regimen, as their weight was below 15 kg. In contrast, 37.9% (95% CI: 37.8–38.2) were excluded based on the height-based approach. This difference was statistically significant (p < 0.001). Figure 5 illustrates the correlation between weight and height among PSAC participants. Panel B of Figure 5 further highlights the lack of concordance between these two criteria, as the intersection of a 15 kg weight and 90 cm height aligns with the 95th percentile. Alternatively, if a fixed-dose of 3 mg were administered to all PSAC without applying any anthropometric criteria for exclusion, the median dose delivered would be 236 µg/kg (IQR: 446 µg/kg). Only 0.26% (95% CI: 0.2–0.3) of children would receive doses exceeding 400 µg/kg, and none would receive doses higher than 600 µg/kg. Figure 5 panel C depicts the median dose achieved by age in PSAC under a fixed 3 mg dosing regimen.

**Figure 5:**
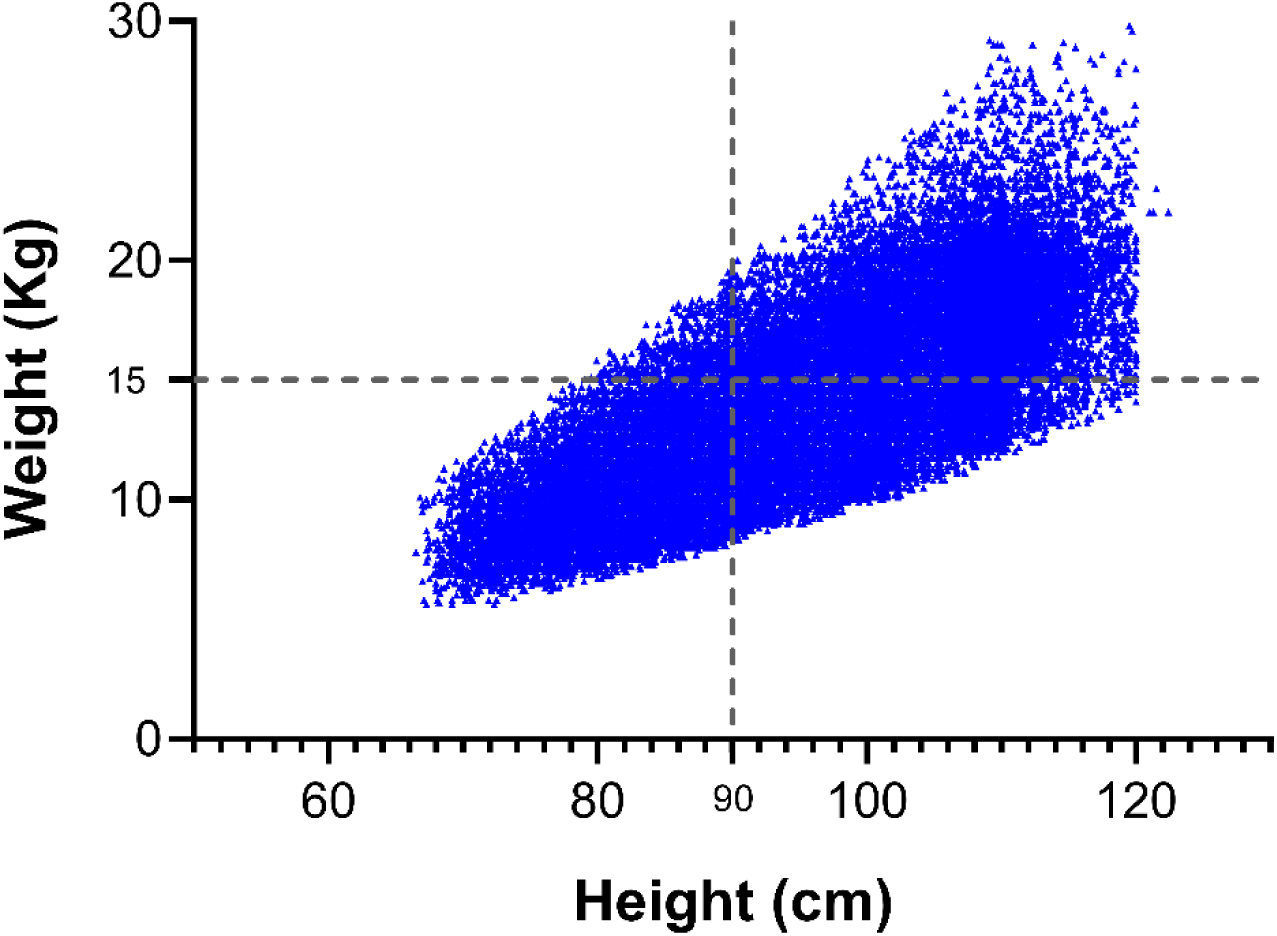
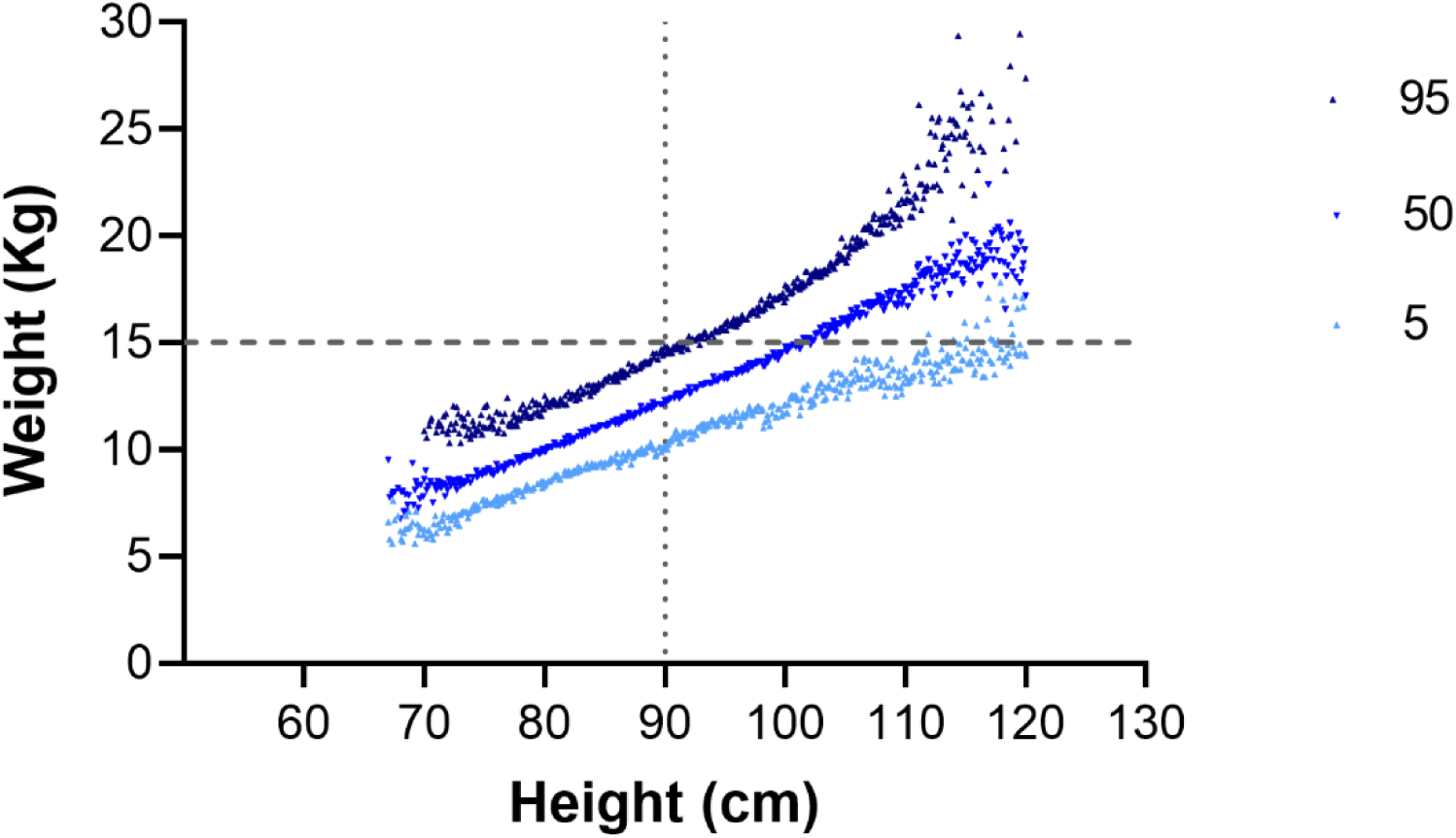
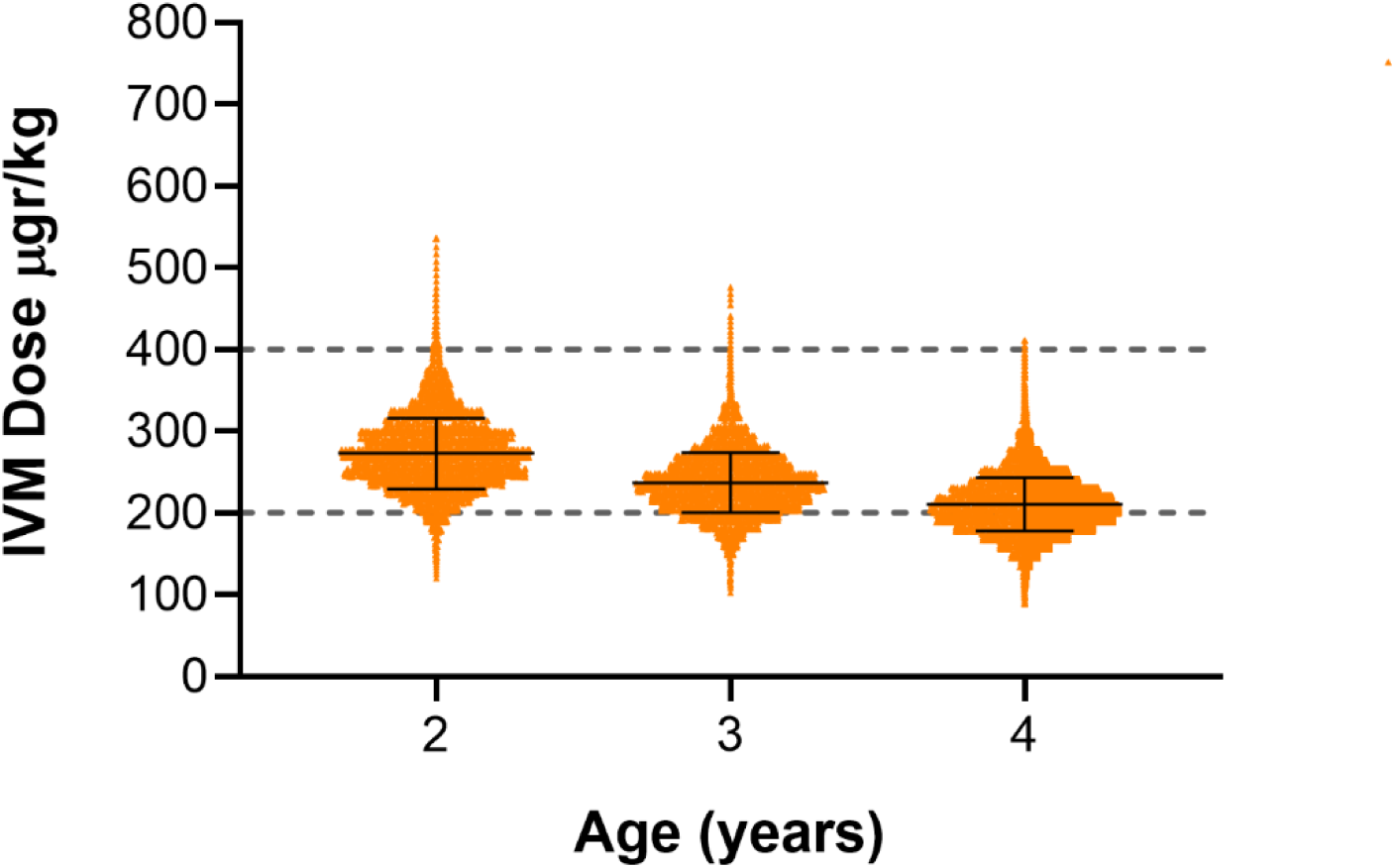
Panel A: Correlation between height and weight of PSAC (N= 317,401); Panel B: weight for height percentiles of PSAC participants of the study; Panel C: Distribution of ivermectin dose (µg/kg) by year of age in PSAC using the fixed-dose regimen. Notes for Figure 5, Panels A and B: Gray dashed lines indicate the current lower limits of the weight-based and height-based recommendations for ivermectin use.

## DISCUSSION

This IPD meta-analysis, which included 741,700 participants from 53 NTD endemic countries, provides a comprehensive assessment of fixed-dose ivermectin regimens. By modeling drug exposure under different dosing strategies and calculating the dose that each participant would receive according to various regimens, our analysis demonstrates that an alternative age-based fixed-dose regimen achieves therapeutic dosing in a higher proportion of individuals compared to weight- and height-based regimens. Furthermore, we found that a simplified dosing approach reduces systematic underdosing without a great risk of exceeding established safety thresholds. This alternative approach, aligned with existing public health classification of at-risk groups (PSAC, SAC and WRA), could simplify drug administration logistics while contributing to dose optimization [1].

Compared to weight- or height-based approaches, an age-based fixed-dose regimen reduces wide-spread underdosing and, significantly increases the proportion of adequately treated individuals, with extremely small risk of doses over 800 µg/kg. Our findings align with those of Goss (2019) regarding IVM underdosing in adult populations and further expand the evidence of this issue in school-aged children (SAC). This concern was also highlighted by Buonfrate (2023), who found that fewer than 50% of SAC participants achieved the optimal dose with the current dosing approaches [10,30].

Available data indicate that IVM doses of 600 mcg/kg, and possibly 800 mcg/kg, are safe and suggest that the target dose range for IVM is due for reconsideration [14–16]. This supports the feasibility of implementing age-based fixed-dose regimens in PSAC, SAC, and adults, ensuring doses remain within the therapeutic index. In adults, trials have demonstrated the safety of single doses up to 2,000 mcg/kg in healthy volunteers and daily doses of 1,200 mcg/kg for five consecutive days in COVID-19 patients [31,32].

PSAC are systematically excluded from IVM treatment due to age, weight, or height criteria. Age-based thresholds exclude children under five, despite surpassing 15 kg [7]. Weight- and height-based regimens lack concordance with growth curves, as their dosing thresholds do not accurately reflect the relationship between height and weight, leading to underdosing or exclusion. Given PSAC’s vulnerability to NTDs, this is concerning. A 3 mg fixed-dose ensures adequate drug exposure while addressing toxicity concerns [12,33].

A key strength of this study is the large number of participants, all from countries endemic for STH and other NTDs. The participants represent "real-world" recipients of MDA interventions, characterized by a high prevalence of malnutrition. As a result, evaluating fixed -dose IVM in this population enhances confidence in the low risk of excessive dosing. It is also relevant the homogeneity of the results across geographic regions.

This study has several limitations. SAC were underrepresented compared to PSAC and WRA, which may affect the generalizability of findings for this group. Despite SAC receiving the highest volume of anthelmintic drugs globally through school-based MDA programs targeting STH, anthropometric data remain scarce. Additionally, determining a fixed dose for SAC was challenging due to weight variation, as growth in this age range is steady. The selected dose prioritized minimizing underdosing.

The study relied on secondary data and the sources of IPD was diverse, which may have introduced inconsistencies or missing values, though heterogeneity analysis showed overall variability was low. Countries were assumed to be similar to be analyzed as a single population, but site-specific factors, such as India’s unique nutritional and regulatory context, warrant further investigation of those sites, beyond the scope of this study.

Findings may also lack applicability to adult males, as no IPD were available for this group. Nonetheless, given that NTDs cause substantial morbidity in men and impact productivity in endemic regions, adult males would also likely benefit from IVM treatment [34,35]. The fixed dose for WRA was set at the upper weight range, ensuring adequate treatment and may also provide appropriate dosing to adult males.

The timing of these findings add urgency to the growing data suggesting that the target dose range for IVM should be revisited. Further research will continue to contribute to our collective understanding, these results offer a strong foundation for considering a shift in practice to a fixed-dose regimen. WHO and national NTD programs may identify specific implementation research needs to assess the cost-effectiveness, and practical application of this approach in endemic settings, alongside its acceptability among healthcare workers and communities. Additionally, pharmaceutical development of paediatric formulations ensuring palatability and bioequivalence for PSAC remains an important research gap [36,37]. The upper safety limit for ivermectin dosing has not yet been clearly established. Further studies, including analyses of data from its widespread use during the COVID-19 pandemic, could help define this limit more precisely [31]. However, any call for additional studies should be weighed against the risk of prolonging the widespread use of sub-therapeutic IVM dosing, which could delay meaningful improvements in treatment outcomes.

In conclusion, the findings of this study provide robust evidence to inform policy discussions on IVM dosing, supporting the feasibility and benefits of transitioning from weight- and height-based IVM dosing to an age-based fixed-dose regimen. They offer critical insights into drug exposure among PSAC, a group currently excluded from MDA interventions. A fixed-dose strategy would reduce the substantial proportion of underdosed individuals without increasing the risk of toxicity [17].

Taken together, these findings present a compelling case as policy-makers at international, regional, and national levels consider updating treatment guidelines. The potential public health benefits -greater efficiency, broader coverage, and improved community engagement-underscore the importance of translating this evidence into action.

## Data Availability

All relevant data are available from the Figshare repository at the following DOI: 10.6084/m9.figshare.28734971.

https://doi.org/10.6084/m9.figshare.28734971

## ACKNOWLEDGEMENTS

We extend our acknowledgment and gratitude to all authors and institutions that contributed their valuable data.

This research includes data obtained through a request to the Infectious Diseases Data Observatory (IDDO) (https://www.iddo.org/data-sharing/accessing-data). IDDO had no role in the production of this research. It also includes data provided by the COVID-19 Data Platform, which is hosted and led by IDDO and the International Severe Acute Respiratory and emerging Infections Consortium (ISARIC), both of which had no role in the production of this research.

Permission to reuse the data was requested by IDDO from the original contributor, or from the Data Access Committee if the responsibility had been delegated by the contributor. This permission was a prerequisite for data sharing. A full list of the organisations and authors that contributed the data in the dataset requested for the present study to the IDDO Data Platform is provided in the Supporting information (S3 Table).

We would like to especially acknowledge Shanti Rochester, Data Governance Officer at the Infectious Diseases Data Observatory (IDDO), Big Data Institute Building, University of Oxford, for her kind assistance during the data sharing process.

We also gratefully acknowledge Professor Bob Taylor and Dr. James Watson for their valuable advice on data collection strategies.

## SUPPORTING INFORMATION

S1 Table: Nutritional assessment by country

S2 Figure: Distribution of IVM dose by age

S3 Table: List of contributors to IDDO dataset

S4 Checklist: PRISMA-IPD Checklist

S5 Table: Ivermectin dosing regimens. Currently recommended wight-based and height-based and alternative age-based fixed-dose.

